# COVID-19 vaccination intention in the UK: Results from the ‘COVID-19 Vaccination Acceptability Study’ (CoVAccS), a nationally representative cross-sectional survey

**DOI:** 10.1101/2020.08.13.20174045

**Authors:** Susan M. Sherman, Louise E. Smith, Julius Sim, Richard Amlôt, Megan Cutts, Hannah Dasch, G James Rubin, Nick Sevdalis

**Author notes:** Sherman and Smith contributed equally to the work and are joint first authors. Corresponding author: Susan M. Sherman. Susan M. Sherman, Senior Lecturer in Psychology. School of Psychology, Keele University, Keele, ST5 5BG, UK. ORCID: https://orcid.org/0000-0001-6708-3398. Louise E. Smith, Post-doctoral Researcher. Department of Psychological Medicine, King’s College London, Weston Education Centre, Cutcombe Road, London, SE5 9RJ, UK. ORCID: https://orcid.org/0000-0002-1277-2564. Julius Sim. Professor of Health Care Research, School of Medicine, Keele University, ST5 5BG, UK. ORCID: https://orcid.org/0000-0002-1816-1676. Richard Amlôt, Head of Behavioural Science in the Emergency Response Department at Public Health England. Porton Down, Salisbury, Wiltshire, SP4 0JG, UK. Megan Cutts, Research Assistant. School of Psychology, Keele University, Keele, ST5 5BG, UK. Hannah Dasch, Research Assistant. Centre for Implementation Science, Health Service and Population Research Department, Institute of Psychiatry, Psychology and Neuroscience, King’s College London, De Crespigny Park, London SE5 8AF, UK. G James Rubin, Reader in the Psychology of Emerging Health Risks. Department of Psychological Medicine, King’s College London, Weston Education Centre, Cutcombe Road, London, SE5 9RJ, UK. ORCID: https://orcid.org/0000-0002-4440-0570. Nick Sevdalis, Professor of Implementation Science and Patient Safety. Centre for Implementation Science, Health Service and Population Research Department, Institute of Psychiatry, Psychology and Neuroscience, King’s College London, De Crespigny Park, London SE5 8AF, UK. ORCID: https://orcid.org/0000-0001-7560-8924.

## Abstract

**Aim:** To investigate factors associated with intention to be vaccinated against COVID-19.

**Methods:** Online cross-sectional survey of 1,500 UK adults, recruited from an existing online research panel. Data were collected between 14^th^ and 17^th^ July 2020. We used linear regression analyses to investigate associations between intention to be vaccinated for COVID-19 “when a vaccine becomes available to you” and socio-demographic factors, previous influenza vaccination, general vaccine attitudes and beliefs, attitudes and beliefs about COVID-19, and attitudes and beliefs about a COVID-19 vaccination.

**Results:** 64% of participants reported being likely to be vaccinated against COVID-19; 27% were unsure and 9% reported being unlikely to be vaccinated. Personal and clinical characteristics, previous influenza vaccination, general vaccination beliefs, and beliefs and attitudes about COVID-19 and a COVID-19 vaccination explained 77% of the variance in vaccination intention. Intention to be vaccinated was associated with more positive general COVID-19 vaccination beliefs and attitudes, weaker beliefs that the vaccination would cause side effects or be unsafe, greater perceived information sufficiency to make an informed decision about COVID-19 vaccination, greater perceived risk of COVID-19 to others but not oneself, older age, and having been vaccinated for influenza last winter (2019/20).

**Conclusions:** Despite uncertainty around the details of a COVID-19 vaccination, most participants reported intending to be vaccinated for COVID-19. Actual uptake will likely be lower. Vaccination intention reflects general vaccine beliefs and attitudes. Campaigns and messaging about a COVID-19 vaccination should emphasize the risk of COVID-19 to others and necessity for everyone to be vaccinated.

## INTRODUCTION

The COVID-19 pandemic has had a huge impact across societies, with governments worldwide imposing restrictions of movement and other measures such as mandatory use of face coverings or quarantine to prevent the spread of the virus. Hopes of returning to normality have been pinned on the availability of a COVID-19 vaccine, and vaccination is central to the UK government’s COVID-19 recovery strategy.(1) Vaccine trials have reported encouraging results indicating that a COVID-19 vaccine is safe and produces a good immune response.(2, 3) However, the success of a vaccination programme will depend on rates of uptake among the population. It is important to prepare and develop effective policies and messaging for vaccination now, in order to maximize uptake when a vaccine becomes available.

There is a wealth of literature investigating factors associated with vaccine uptake. Research is underpinned by multiple theories of health behaviour, including the Health Belief Model,(4) the Theory of Planned Behaviour,(5) and Protection Motivation Theory.(6) Constructs outlined by these theories, including threat appraisal, coping appraisal, cues to action, self-efficacy, perceived benefits and barriers, subjective norms, perceived behavioural control, and attitudes, have consistently been associated with uptake of routine vaccination (7, 8) as well as vaccine uptake during the H1N1 influenza pandemic.(9) In addition to these theoretical constructs, contextual factors are also known to affect vaccine uptake.(7) Perceptions and attitudes are in part driven by contextual factors, such as current events in the news and how the vaccine is being portrayed in the media.

To date, there have been two studies to our knowledge investigating factors associated with intention to be vaccinated against COVID-19 in the UK in clinically vulnerable populations.(10, 11) One study found that increased intention to be vaccinated was associated with thinking that the COVID-19 outbreak would last for a long time, while decreased intention was associated with thinking that the risks of COVID-19 have been exaggerated by the media.(10) These results should be interpreted cautiously as they did not account for the influence of participants’ socio-demographic characteristics. The second study investigated associations between vaccine intention and sociodemographic factors, finding that decreased intention was associated with younger age and Black and minority ethnicity, but did not investigate the influence of psychological factors on vaccination intention.(11) Results from these studies should be interpreted with caution due to the protracted nature of data collection in both studies and the analyses used. However, they provide some initial insight into factors associated with COVID-19 vaccination intention. It is likely that a COVID-19 vaccination will become available first to those in at-risk groups and those who have increased exposure to the virus through their job.(12) However, vaccination intention in the general population should be investigated as vaccination may be rolled out more widely soon afterwards, and sufficient uptake will be critical to eliminating COVID-19. Furthermore, it remains a possibility that people may be able to be vaccinated for COVID-19 privately, much like the seasonal influenza vaccine.

The aim of this study was to investigate associations between vaccination intention and theoretically-grounded, contextual and socio-demographic factors in a demographically-representative sample of the UK adult population.

## METHOD

### Design

We conducted a cross-sectional survey, between 14^th^ and 17^th^ July 2020. Participants completed the survey online, on Qualtrics.

### Participants

Participants (n=1,500) were recruited through Prolific’s online research panel and were eligible for the study if they were aged eighteen years or over and lived in the UK (n=38,000+ eligible participants). Quota sampling was used, based on age, sex, and ethnicity to ensure that the sample was broadly representative of the UK general population. Of 1,532 people who began the survey, 1,504 completed it (98% completion rate). Four participants were not included in the sample as they did not meet quality control checks. Participants were paid £2 for a completed survey.

### Study materials

Full survey materials are available online.(13) Items were based on previous literature.(14-18)

#### Outcome measures

To measure vaccination intention, we asked participants to state how likely they would be to have a COVID-19 vaccination “when a coronavirus vaccination becomes available to [them]” on an eleven-point scale from “extremely unlikely” (0) to “extremely likely” (10).

#### Psychological factors

We asked participants to what extent they thought “coronavirus poses a risk to” people in the UK and to themselves personally, on a five-point scale, from “no risk at all” to “major risk”.

We asked participants if they thought they “have had, or currently have, coronavirus”. Participants could answer “I have definitely had it or definitely have it now”, “I have probably had it or probably have it now”, “I have probably not had it and probably don’t have it now”, and “I have definitely not had it and definitely don’t have it now”. We also asked participants if they personally knew anyone who had had COVID-19 (yes/no).

Participants were asked a series of statements about COVID-19 (n=8) and about a possible COVID-19 vaccination (n=24). For questions about the COVID-19 vaccination, participants were asked to imagine that a COVID-19 vaccine was widely available. Statements measured theoretical constructs including perceived susceptibility to COVID-19, severity of COVID-19 benefits of a COVID-19 vaccine, barriers to being vaccinated against COVID-19, ability to be vaccinated (self-efficacy), subjective norms, behavioural control, anticipated regret, knowledge, trust in the Government and NHS. These items also investigated concerns about commercial profiteering, and participants’ beliefs about vaccination allowing life to get back to ‘normal’ and having to follow social distancing and other restrictions for COVID-19 if vaccinated. Participants rated perception statements on an eleven-point scale (0–10) from “strongly disagree” to “strongly agree”. We also asked participants if their employer would want them to have the COVID-19 vaccination. Order of items was quasi-randomized.

#### Personal and clinical characteristics

We asked participants to report their age, gender, ethnicity, religion, employment status, highest educational or professional qualification, and total household income. We also asked participants what UK region they lived in, how many people lived in their household, whether they or someone else in their household (if applicable) had a chronic illness that made them clinically vulnerable to serious illness from COVID-19, and if they worked or volunteered in roles considered critical to the COVID-19 response (‘key worker’ roles).

Lastly, we asked participants if they had been vaccinated for seasonal influenza last winter (yes/no), and how likely they would be to have the seasonal influenza vaccine this winter (eleven point scale, from “extremely unlikely” to “extremely likely”).

### Ethics

Ethical approval for this study was granted by Keele University’s Research Ethics Committee (reference: PS-200129).

### Patient and public involvement

Due to the rapid nature of this research, the public was not involved in the development of the survey materials.

### Power

A target sample size of 1500 was chosen to provide a high ratio of cases to estimated parameters in order to avoid overfitting and loss of generalizability in the regression model.(19)

### Analysis

To identify variables associated with an intention to have the COVID-19 vaccination, we constructed a linear regression model. Ordinal and multinomial predictors were converted to dummy variables. To aid interpretation of the model, and to address collinearity in some variables, we ran principal component analyses on items investigating beliefs and attitudes about COVID-19 and a COVID-19 vaccination.

Variables entered into the model were selected a priori based on their theoretical relevance; no variable selection procedures were employed. Five groups of variables were included in the model: personal and clinical characteristics; seasonal influenza vaccination; general beliefs and attitudes relating to vaccination; beliefs and attitudes relating to COVID-19 illness; and beliefs and attitudes relating to COVID-19 vaccination. The percentage of variance in the outcome variable explained by each predictor was calculated as the squared semi-partial correlation for a numerical predictor and the change in *R*^2^ attributable to a set of dummy variables.

As well as fitting the full model, we also added the groups of variables as successive blocks in a hierarchical model, to determine the incremental increase in the adjusted *R*^2^ value as the groups of variables were added to the model.

Due to the large number of predictors in the model, statistical significance was set at *p*≤.01 to control for Type 1 errors and 99% confidence intervals (CIs) were correspondingly calculated for the regression coefficients. Assumptions of the analysis were checked. Analyses were conducted in SPSS 26.

## RESULTS

As intended, participants were broadly representative of the UK population (mean age 46.0 years, SD=15.8, range 18 to 87; 51% female; 85% white ethnicity; Table 1, see supplementary materials 1 for further breakdown).

**Table 1.**
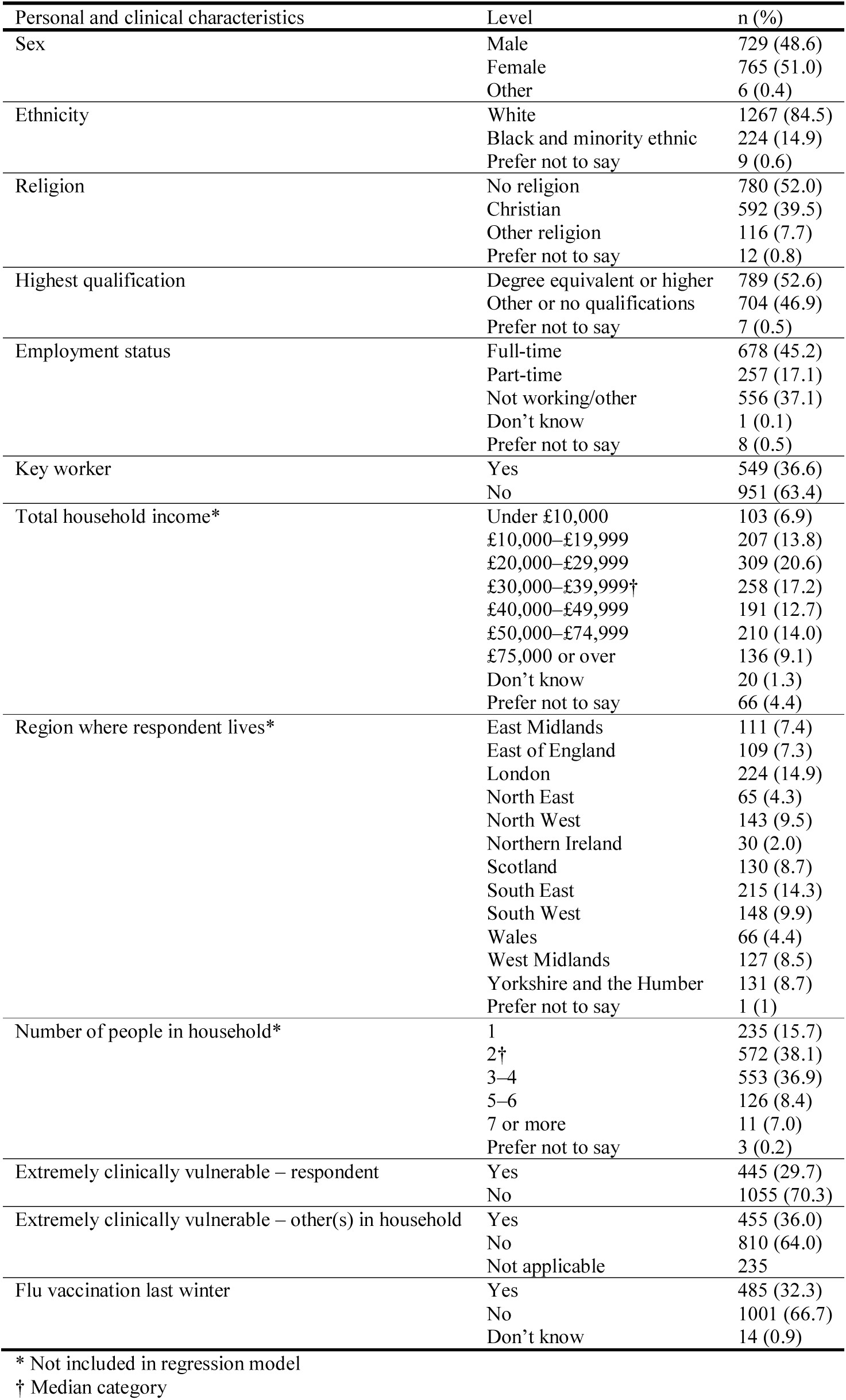
Participant characteristics.

**Table 2.**
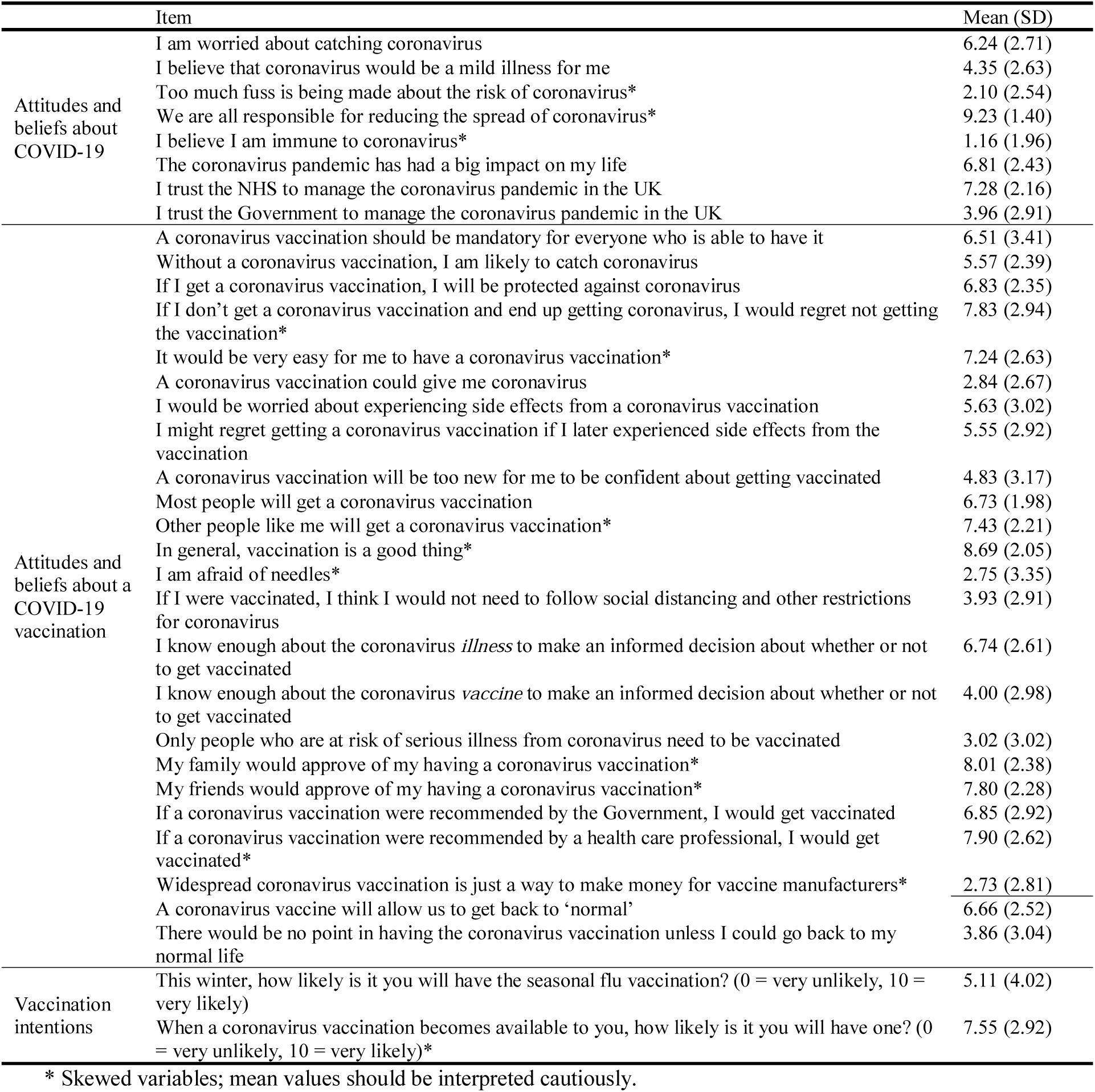
Descriptive statistics for continuous items measuring beliefs and attitudes about COVID-19 and a COVID-19 vaccination and vaccination intention. Data are mean (standard deviation) on a 0-10 numerical rating scale (0 = strongly disagree, 10 = strongly agree).

### Principal component analyses

Two components emerged from the principal component analysis on beliefs and attitudes about COVID-19 (see supplementary materials 2). The first component reflected items about perceived severity of COVID-19, perceived immunity to the virus, and impact of the pandemic on one’s life (*“perceived threat and impact of COVID-19’*). The second component measured trust in the NHS and the Government to manage the COVID-19 pandemic in the UK (*“trust in COVID-19 management”*).

When investigating items related to a COVID-19 vaccination, four components emerged from this principal component analysis (see supplementary materials 2). The first component measured *“general COVID-19 vaccination beliefs and attitudes”*, with items loading onto this factor investigating perceived vaccine effectiveness, social norms, likelihood of catching COVID-19 without a vaccine, beliefs about mandatory vaccination, the influence of vaccine recommendations from different sources, anticipated regret of not being vaccinated, and perceived ease of vaccination. The second component, termed *“COVID-19 vaccination adverse effects”* measured perception of adverse effects and novelty of the vaccine. The third component measured perceived information sufficiency to be able to make an informed decision about vaccination (*“perceived knowledge sufficiency*”). Items about vaccination allowing a return to ‘normal’ life and not having to follow social distancing and other restrictions if one were vaccinated loaded on to the fourth component (*return to ‘normal ’ life*).

### Vaccination intention

Participants’ vaccination intention is presented in Figure 1. Vaccination intention exhibited a marked negative skew (mean=7.55, standard deviation=2.92, median=9). Using a priori categorisations (scores of zero to two as “very unlikely”, three to seven as “uncertain” and eight to ten as “very likely”), 9.1% (95% CI 7.8%, 10.7%) reported being very unlikely to be vaccinated (*n*=137), 26.9% (95% CI 24.7%, 29.2%) reported being uncertain about their likelihood of vaccination (*n*=403), and 64.0% (95% CI 61.5%, 66.4%) reported being very likely to be vaccinated (*n*=960).

**Figure 1.**
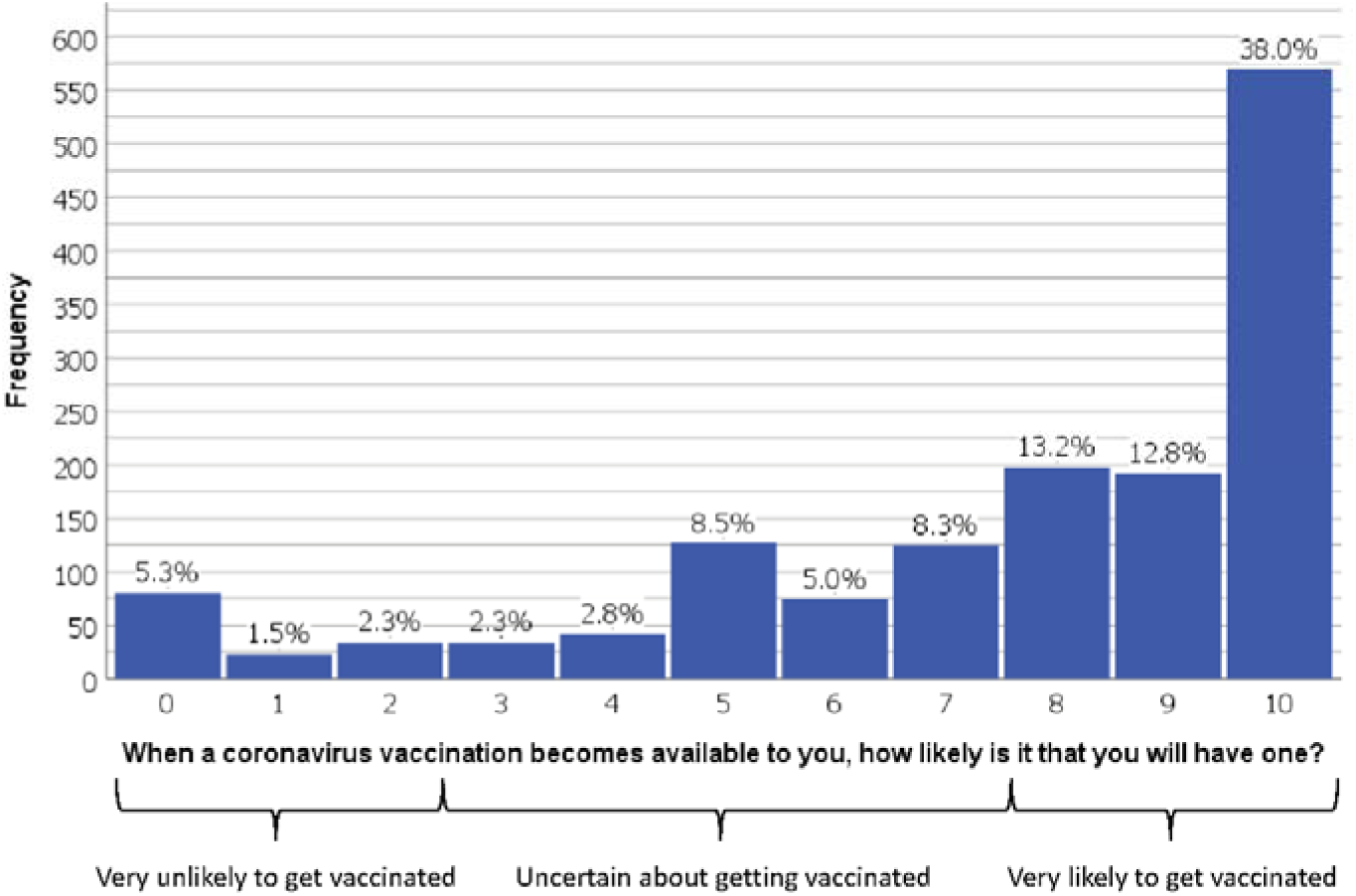
Perceived likelihood of having a vaccination (0=“extremely unlikely” to 10=“extremely likely”.)

The final model explained 77% of the variance in intention to vaccinate (Table 3). Increased likelihood of being vaccinated for COVID-19 was significantly associated with older age, having been vaccinated for influenza last winter, perceiving a greater risk of COVID-19 to people in the UK, more positive general COVID-19 vaccination beliefs and attitudes, weaker beliefs that the vaccination would cause side effects or be unsafe, greater perceived information sufficiency to make an informed decision about COVID-19 vaccination, and lower endorsement of the notion that only people who are at risk of serious illness should be vaccinated for COVID-19. In terms of explained variance, the strongest predictors were the principal components representing general COVID-19 vaccination beliefs and attitudes (19.5% of variance explained), followed by vaccination adverse effects and novelty (8.2% of variance explained).

**Table 3.** Descriptive statistics for categorical and ordinal items measuring beliefs and attitudes about COVID-19 and a COVID-19 vaccination.

| Item | Level | n (%) |
| --- | --- | --- |
| To what extent do you think coronavirus poses a risk to people in the UK? | No risk at all<br>Minor risk<br>Moderate risk<br>Significant risk<br>Major risk<br>Don't know | 5 (0.3)<br>80 (5.3)<br>313 (20.9)<br>675 (45.0)<br>426 (28.4)<br>1 (0.1) |
| To what extent do you think coronavirus poses a risk to you personally? | No risk at all<br>Minor risk<br>Moderate risk<br>Significant risk<br>Major risk<br>Don't know | 34 (2.3)<br>365 (24.3)<br>563 (37.5)<br>381 (25.4)<br>153 (10.2)<br>4 (0.3) |
| Do you believe you have had, or currently have, coronavirus? | Definitely not<br>Probably not<br>Probably<br>Definitely<br>Don't know | 555 (37.0)<br>588 (39.2)<br>151 (10.1)<br>28 (1.9)<br>178 (11.9) |
| Do you personally know anyone (excluding yourself) who has had coronavirus? | Yes<br>Don't know<br>Prefer not to say | 677 (45.2)<br>822 (54.8)<br>1 (0.1) |
| As far as you know, would your employer want you to have the coronavirus vaccination? | Yes<br>No<br>Don't know<br>Not applicable | 597 (61.2)<br>18 (1.8)<br>361 (37.0)<br>524 |
| As far as you know, is there currently a widely available vaccination to protect against coronavirus? | Yes<br>No<br>Don't know<br>Prefer not to say | 53 (3.5)<br>1360 (90.7)<br>82 (5.5)<br>5 (0.3) |

**Table 3.**
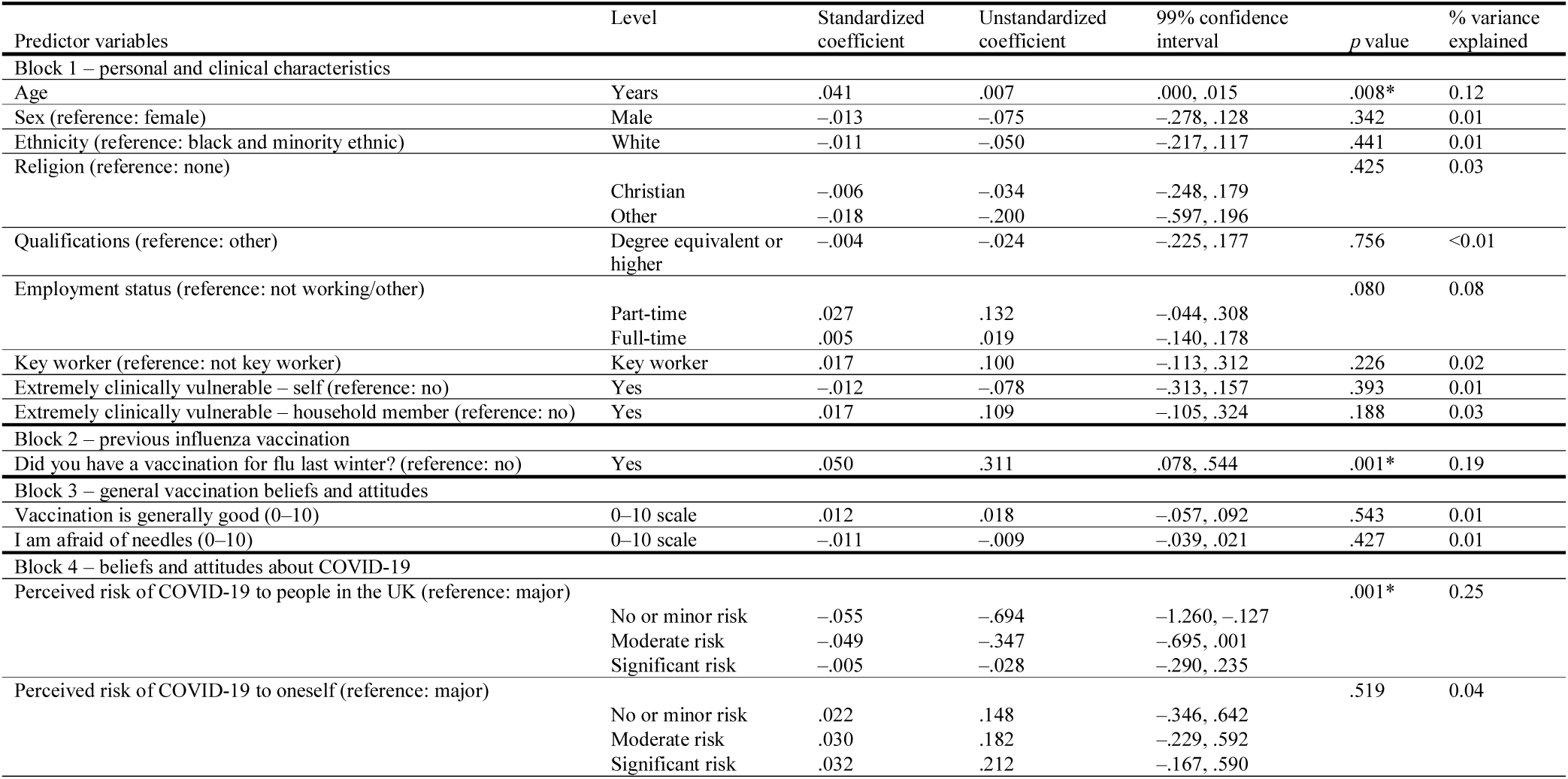
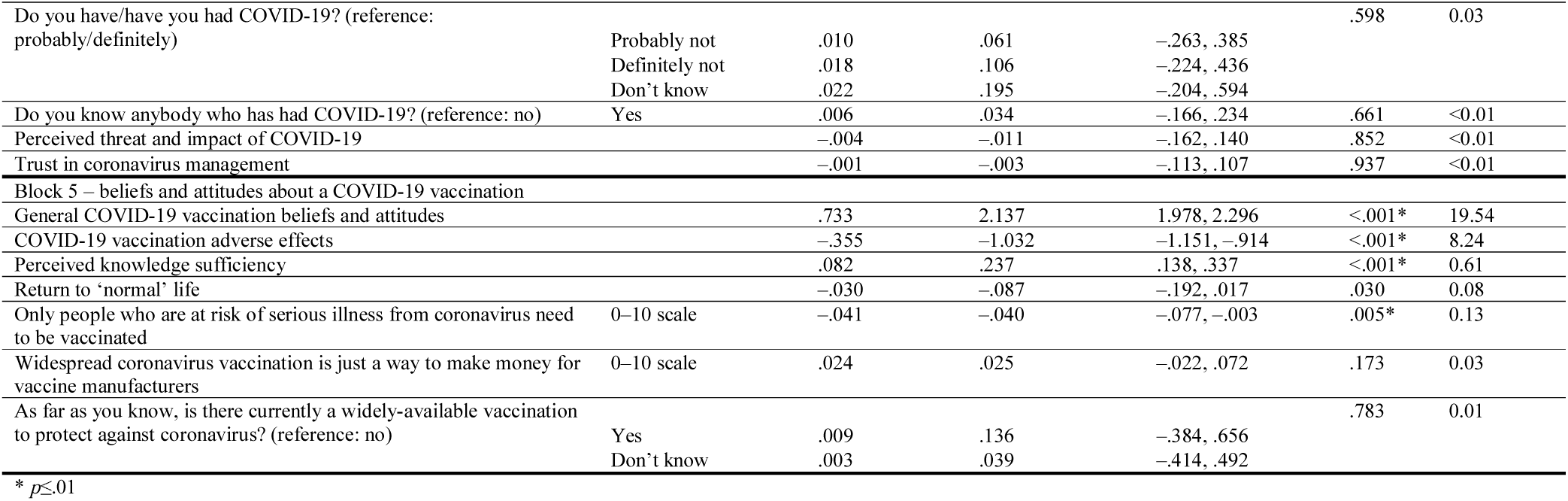
Results of the full linear regression model analysing associations with vaccination intention (adjusted *R*^2^ = .771). Parameter estimates relate to the full model containing all predictors. The unstandardized regression coefficients represent the change in likelihood of vaccination for a one-unit increase in the predictor variable (or, for dummy variables, a shift from the reference category to the category concerned). The figures under ‘% variance explained’ represent the percentage of variance in the outcome variable uniquely explained by the item (or set of dummy variables) concerned. The model was based on 1448 cases with complete data.

When the groups of variables were entered hierarchically as blocks, we can infer the percentage of additional variance explained by each block from the change in incremental adjusted *R*^2^. Addition of each subsequent block explained a statistically significant proportion of the variance (*p*<.001 in each case). Personal and clinical characteristics (block 1) alone explained very little (3%) of the variance in intention to be vaccinated. When previous influenza vaccination (block 2) was added, it explained an additional 7% of the variance. Adding general vaccination beliefs and attitudes (block 3) resulted in the largest increase in proportion (35%) of explained variance (though in the full model the predictors in this group were no longer significant). When beliefs and attitudes about COVID-19 (block 4) were added to the model, they explained 4% more of the variance in vaccination intention. Adding positive beliefs and attitudes about a COVID-19 vaccination (block 5) explained a further 28% of the variance.

## DISCUSSION

If COVID-19 vaccination were to be offered to the general population, one advantage might be the ability to achieve herd immunity. Estimates indicate that up to 60% of the population might need to be vaccinated to achieve this.(20) Sixty-four percent of people surveyed reported intending to be vaccinated for COVID-19 when a vaccine becomes available to them. While intention is a key driver of the uptake of health behaviours,(5, 6) vaccination intention is likely to be higher than actual vaccine uptake.(21) Therefore, it is important to identify factors associated with vaccination intention early on, to support policy and communications when a vaccine becomes available. We found that, taken together, personal and clinical characteristics, previous influenza vaccination, general vaccination beliefs, and beliefs and attitudes about COVID-19 and a COVID-19 vaccination explained 77% of the variance in vaccination intention.

Importantly, we found that the factor that explained the greatest proportion of the variance in vaccination intention (20%) was COVID-19 vaccination beliefs and attitudes. This factor encompassed items measuring positive influence of recommendations from authorities to be vaccinated, greater perceived social norms about vaccination, greater perceived effectiveness, greater perceived likelihood of catching COVID-19 without a vaccine, greater anticipated regret of not being vaccinated, beliefs that COVID-19 vaccination should be mandatory and greater perceived ease of vaccination. These map on to theoretical constructs of threat appraisal, coping appraisal, subjective norms, and self-efficacy outlined by theories of uptake of health behaviours,(4-6) which were also associated with uptake of vaccination during the H1N1 influenza pandemic.(9) Earlier research investigating COVID-19 vaccine willingness in the UK found no association with perceived likelihood of catching COVID-19, trust in authorities, or clarity of information about the virus.(10) However, those earlier results should be interpreted with caution as analyses did not control for personal or clinical characteristics and data were collected early in the pandemic. Our results suggest that people may hold general positive or negative beliefs and attitudes towards the vaccination and this general sense is driving vaccination intention at this point. While a COVID-19 vaccination has so far been positively framed in the media, as more information - and misinformation - about the vaccine comes to light, there is the potential for this general positive sentiment to be eroded, negatively influencing vaccination intention and uptake.

Details around COVID-19 vaccination remain uncertain until a vaccine has been developed, but will become clearer as more information is available regarding vaccine composition (immunogenicity and safety), and immunity after having contracted COVID-19.(12) We found that vaccination intention was associated with greater perceived information sufficiency about COVID-19 and a COVID-19 vaccination. In the case of COVID-19, a perception of sufficient information about the vaccination is interesting as there is currently little information available about a vaccine. What information there is comes from results of vaccine trials that were still underway at the time of data collection. These results may therefore reflect participants’ general vaccine beliefs and attitudes.

In contrast to previous research,(9) we found no evidence of an association between greater perceived risk of COVID-19 to oneself and vaccination intention. However, greater perceived risk to others was associated with vaccination intention in our study. This suggests that vaccination campaigns and messaging highlighting the need for vaccination for altruistic reasons (i.e. to protect others) might be particularly effective. We also found that concerns about adverse effects and vaccine novelty were associated with vaccination intention. As novel threats are perceived as inherently more risky,(22) and perceiving adverse effects is consistently associated with vaccination refusal,(8, 9) this is unsurprising.

Eligibility criteria for a COVID-19 vaccination are not yet clear. Initial guidance from the Joint Committee on Vaccination and Immunisation suggests that vaccination should be prioritized among frontline health and social care workers and those at increased risk of critical illness or death from COVID-19.(12) We found no evidence of an association between clinical vulnerability to COVID-19 and vaccination intention. However, vaccination intention was lower in those who thought that only those who are at risk of serious illness need to be vaccinated. This may be because most of the sample did not think that they were at increased clinical risk of COVID-19. Our findings that thinking that one has had COVID-19 was not associated with vaccination intention is reassuring.

With some evidence suggesting that repeated vaccination for COVID-19 may be necessary,(20) parallels with seasonal influenza vaccination can be drawn and lessons learned to promote vaccination uptake. Populations at greater clinical risk of serious illness from COVID-19 are also similar to those at-risk of serious illness from influenza, and target populations for vaccines are likely to be similar. We found that seasonal influenza vaccination was strongly associated with COVID-19 vaccination intention. With the 2020/21 influenza season fast approaching in the UK, and an increasing strain that concurrent circulation of seasonal influenza and COVID-19 will put on healthcare services,(23) it is crucial that uptake of the seasonal influenza vaccine increases compared to uptake in 2019/20 (England: 72% in 65+ year olds, 45% in a clinical risk category; 44% in pregnant women; and 44% in pre-school children and 60% in school-aged children (24)).

Given the prominence of COVID-19 in the media, contextual factors are likely to be strongly influential in vaccination uptake,(7) with vaccine sentiments likely reflecting the media discourse. However, there was no evidence for an association between beliefs about a return to ‘normal’ and COVID-19 vaccination intention using our stringent criteria for statistical significance (*p*≤ .01). This may be due to the continuing uncertainties surrounding a COVID-19 vaccination. Given the potential for sensationalized stories to increase perceptions of the likelihood and severity of adverse effects, decrease vaccine uptake, and in some cases lead to political responses including the suspension of vaccination programmes,(25) it is important that when more information about a vaccine becomes available, a clear factual account is portrayed in the media. It remains to be seen how this might be implemented in practice.

In line with other research conducted on COVID-19 vaccine willingness in the UK,(11) we found that older age was associated with greater intention to be vaccinated. This finding may reflect the related increased uptake of seasonal influenza vaccination in older age groups.

This study has limitations. First, although we used a demographically representative sample of the UK population, we cannot be sure how representative survey respondents are of the views and behaviours of the general population.(26, 27) However, we assume that associations between variables follow the same pattern as those in the general population.(28)

Second, we cannot infer causality due to the cross-sectional nature of the study. Third, we investigated vaccination intention. Actual vaccination uptake is likely to be lower. (21) Given the theoretical importance of intention in theories of uptake of health behaviours,(5, 6) it is likely that factors associated with vaccination intention in this study will also influence vaccination uptake. Fourth, due to unclear evidence of the role of children in transmission of COVID-19 in the UK (12) and space constraints in the survey, we chose not to investigate intention to vaccinate one’s child for COVID-19.

High levels of uptake of a COVID-19 vaccination when one becomes available will be necessary in order for the UK government’s COVID-19 recovery strategy to be fulfilled and for life to return to ‘normal’. To the best of our knowledge, this is the first methodologically rigorous study investigating intention to receive a COVID-19 vaccination in a demographically-representative sample of the UK population. While there is still much uncertainty surrounding COVID-19 and vaccination, results from this study provide useful insights that can help guide policy and communications when a vaccine becomes available. The UK population is still divided in their intention to be vaccinated for COVID-19. Approximately two-thirds report being likely to be vaccinated when a vaccine becomes available to them despite the dearth of information about a COVID-19 vaccination. As vaccine uptake is likely to be lower than vaccination intention, it is worrying that the remaining third were unsure or did not intend to be vaccinated for COVID-19, given the impact of COVID-19 on day-to-day life and prominence of the virus in the media. These findings are likely to reflect general vaccine attitudes and beliefs. Our results indicate that vaccination campaigns and communications should draw on theoretical constructs. Contextual factors, such as the media discourse around a COVID-19 vaccination, are likely to influence beliefs and attitudes towards the vaccine. Communications should also explain and highlight how vaccination can stop the spread of COVID-19 to others and facilitate a return to normality.

## Data Availability

The link provided is a read-only link. On acceptance this will be updated to a fully accessible link to the data.

https://osf.io/94856/?view_only=c85fd2666a204c67b2c41f0ded105ec2

## TRANSPARENCY DECLARATION

The authors affirm that the manuscript is an honest, accurate, and transparent account of the study being reported; that no important aspects of the study have been omitted; and that any discrepancies from the study as originally planned have been explained.

## DATA SHARING STATEMENT

Data are available online.(13)

## AUTHOR CONTRIBUTION STATEMENT

The study was conceptualized by SS, LS, JS, RA, GJR and NS. Materials were developed by SS, LS, JS, MC, HD, GJR and NS. SS and MC completed data cleaning. JS completed analyses. LS wrote the first draft of the manuscript. All authors contributed to, and approved, the final manuscript.

## SUPPLEMENTARY MATERIALS 1

Table showing full breakdown of participant characteristics. Data are frequencies (%) except where indicated.

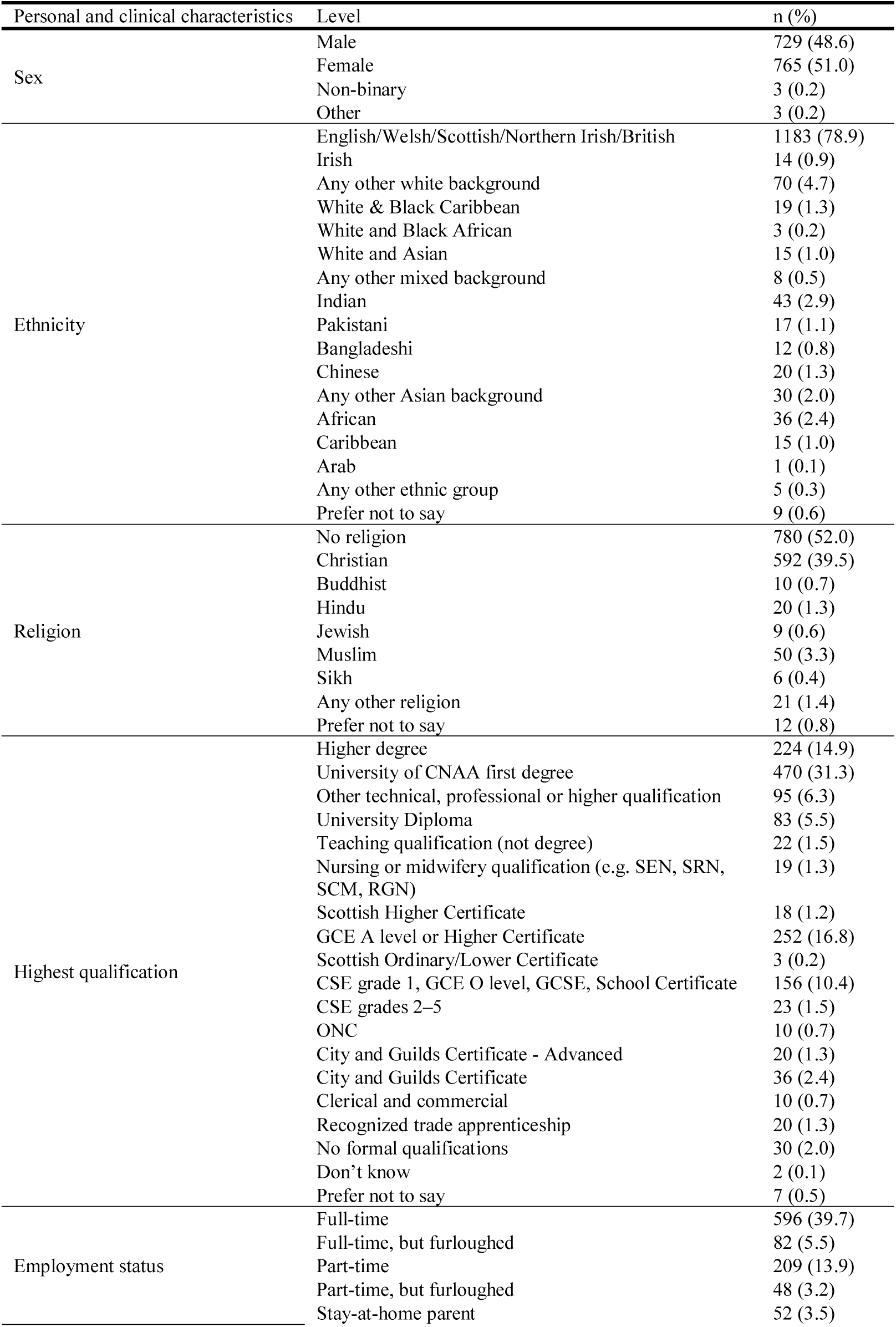
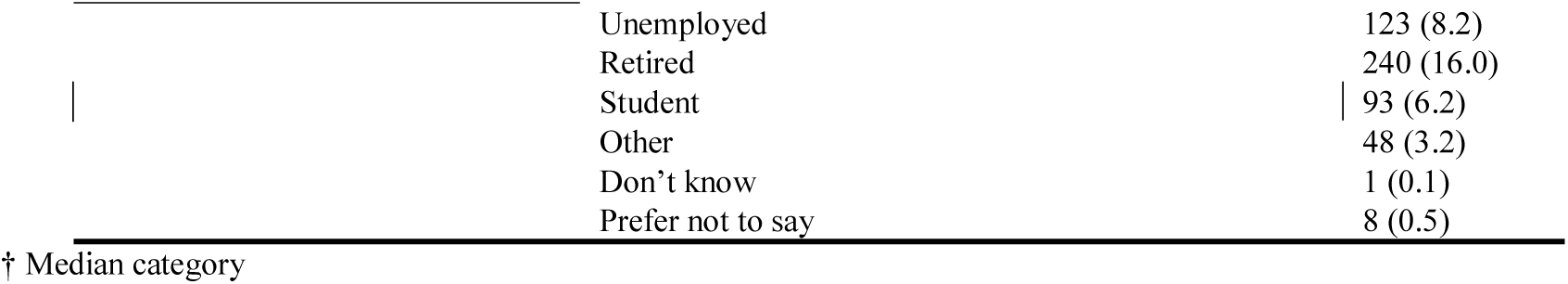

## SUPPLEMENTARY MATERIALS 2

Table showing principal component loadings, following varimax rotation for items relating to attitudes and beliefs about COVID-19. Only loadings over .400 are shown.

|  | Perceived threat and impact of<br>COVID-19 | Trust in management of<br>COVID-19 |
| --- | --- | --- |
| I am worried about catching coronavirus | -.807 |  |
| I believe that coronavirus would be a mild illness<br>for me | .652 |  |
| Too much fuss is being made about the risk of<br>coronavirus | .757 |  |
| We are all responsible for reducing the spread of<br>the coronavirus | -.585 |  |
| I believe I am immune to coronavirus | .564 |  |
| The coronavirus pandemic has had a big impact on<br>my life | -.454 |  |
| I trust the NHS to manage the coronavirus<br>pandemic in the UK |  | .810 |
| I trust the Government to manage the coronavirus<br>pandemic in the UK |  | .778 |

Table showing principal component loadings, following varimax rotation for items relating to attitudes and beliefs about a COVID-19 vaccination. Only loadings over .400 are shown.

|  | General COVID-19<br>vaccination beliefs and<br>attitudes | Vaccination<br>adverse effects | Perceived<br>knowledge<br>sufficiency | Return to<br>'normal'<br>life |
| --- | --- | --- | --- | --- |
| A coronavirus vaccination should be mandatory for everyone who is able to have it | 0.684 |  |  |  |
| Without a coronavirus vaccine, I am likely to catch coronavirus | 0.633 |  |  |  |
| If I get a coronavirus vaccination, I will be protected against coronavirus | 0.716 |  |  |  |
| If I don't get a coronavirus vaccination and end up getting coronavirus, I would regret not getting the vaccination | 0.739 |  |  |  |
| It would be very easy for me to have a coronavirus vaccination | 0.585 |  |  |  |
| A coronavirus vaccination could give me coronavirus |  | 0.629 |  |  |
| I would be worried about experiencing side effects from a coronavirus vaccination |  | 0.809 |  |  |
| I might regret getting a coronavirus vaccination if I later experienced side effects from the vaccination |  | 0.733 |  |  |
| A coronavirus vaccination will be too new for me to be confident about getting vaccinated |  | 0.696 |  |  |
| Most people will get a coronavirus vaccination | 0.594 |  |  |  |
| Other people like me will get a coronavirus vaccination | 0.759 |  |  |  |
| If I were vaccinated, I think I would not need to follow social distancing and other restrictions for coronavirus |  |  |  | 0.821 |
| I know enough about the coronavirus illness to make an informed decision about whether or not to get vaccinated |  |  | 0.717 |  |
| I know enough about the coronavirus vaccine to make an informed decision about whether or not to get vaccinated |  |  | 0.844 |  |
| My family would approve of my having a coronavirus vaccination | 0.775 |  |  |  |
| My friends would approve of my having a coronavirus vaccination | 0.755 |  |  |  |
| If a coronavirus vaccination were recommended by the Government, I would get vaccinated | 0.756 |  |  |  |
| If a coronavirus vaccination were recommended by a health care professional (e.g. GP or nurse), I would get vaccinated | 0.803 |  |  |  |
| A coronavirus vaccine will allow us to get back to 'normal' | 0.663 |  |  | 0.417 |
| There would be no point in having the coronavirus vaccination unless I could go back to my normal life |  |  |  | 0.655 |

